# Efficacy of hydroxychloroquine in patients with COVID-19: results of a randomized clinical trial

**DOI:** 10.1101/2020.03.22.20040758

**Authors:** Zhaowei Chen, Jijia Hu, Zongwei Zhang, Shan Jiang, Shoumeng Han, Dandan Yan, Ruhong Zhuang, Ben Hu, Zhan Zhang

## Abstract

**Aims:** Studies have indicated that chloroquine (CQ) shows antagonism against COVID-19 in vitro. However, evidence regarding its effects in patients is limited. This study aims to evaluate the efficacy of hydroxychloroquine (HCQ) in the treatment of patients with COVID-19.

**Main methods:** From February 4 to February 28, 2020, 62 patients suffering from COVID-19 were diagnosed and admitted to Renmin Hospital of Wuhan University. All participants were randomized in a parallel-group trial, 31 patients were assigned to receive an additional 5-day HCQ (400 mg/d) treatment, Time to clinical recovery (TTCR), clinical characteristics, and radiological results were assessed at baseline and 5 days after treatment to evaluate the effect of HCQ.

**Key findings:** For the 62 COVID-19 patients, 46.8% (29 of 62) were male and 53.2% (33 of 62) were female, the mean age was 44.7 (15.3) years. No difference in the age and sex distribution between the control group and the HCQ group. But for TTCR, the body temperature recovery time and the cough remission time were significantly shortened in the HCQ treatment group. Besides, a larger proportion of patients with improved pneumonia in the HCQ treatment group (80.6%, 25 of 31) compared with the control group (54.8%, 17 of 31). Notably, all 4 patients progressed to severe illness that occurred in the control group. However, there were 2 patients with mild adverse reactions in the HCQ treatment group. Significance: Among patients with COVID-19, the use of HCQ could significantly shorten TTCR and promote the absorption of pneumonia.

**Significance:** Among patients with COVID-19, the use of HCQ could significantly shorten TTCR and promote the absorption of pneumonia.

**Trial registration:** URL: https://www.clinicaltrials.gov/. The unique identifier: ChiCTR2000029559.

## Introduction

Coronaviruses are enveloped positive-sense single-stranded RNA viruses belonging to the family Coronaviridae and are broadly distributed in humans and other vertebrates, eventually causing damage in digestive, respiratory and even multiple systems. In December 2019, a series of pneumonia cases of unknown etiology appeared in Wuhan, Hubei, China [1]. Sequencing analysis of throat swabs samples and electron microscope observations indicated a novel coronavirus, which was named SARS-CoV-2 (formerly known as 2019-nCoV) [2]. Coronavirus disease 2019 (COVID-19) caused by SARS-CoV-2 has been confirmed to have obvious human-to-human characteristics [3,4]. As of March 20, 2020, more than two hundred thousand confirmed cases have been identified globally, for a total of 8778 deaths [5]. As the epidemic is spreading to many countries, COVID-19 poses a severe threat to global health [6]. Therefore, it is urgent to develop effective drugs against COVID-19.

The recent publication of results showing the activity of chloroquine (CQ) against SARS-CoV-2 *in vitro* [7], some experts and researchers also have been recommended the efficacy of this antimalarial drug in patients with COVID-19 [8,9]. For this, the U.S. Food and Drug Administration (FDA) has been working to investigate the use of CQ in COVID-19 [10]. As a derivative of CQ, hydroxychloroquine (HCQ) has similar therapeutic effects and fewer adverse effects. Based on its characteristics of immunity regulation, antithrombotic activity, and inflammation improvement, HCQ has been routinely used in the clinical treatment of systemic lupus erythematosus (SLE) [11]. However, the efficacy of HCQ in COVID-19 remains unknown.

Interestingly, through a follow-up survey, we found that none of our 80 SLE patients who took long-term oral HCQ had been confirmed to have SARS-CoV-2 infection or appeared to have related symptoms. In addition, among the 178 patients diagnosed with COVID-19 pneumonia in our hospital, none were receiving HCQ treatment before admission. All predicting the use of HCQ in SARS-CoV-2 infections. As one of the clinical research registration units in China, we aimed to investigate the efficiency of HCQ in patients with COVID-19 in this study.

## Materials and Methods

### Study design and participants

The clinical research protocol was reviewed and approved by the Ethics Committee in Renmin Hospital of Wuhan University (Wuhan, China). All research procedures adhered to the tenets of the Declaration of Helsinki. This trial for SARS-CoV-2 has already been registered in the Chinese Clinical Trial Registry (ChiCTR), the unique identifier: ChiCTR2000029559. Informed consent was obtained from all patients.

From February 4, 2020, to February 28, 2020, 142 patients with confirmed COVID-19 were admitted. Diagnosis and classification of COVID-19 were based on the criteria of the China National Health Commission. For this trial, the selection criteria: 1. Age ≥ 18 years; 2. Laboratory (RT-PCR) positive of SARS-CoV-2; 3. Chest CT with pneumonia; 4. SaO_2_/SPO_2_ ratio > 93% or PaO_2_/FIO_2_ ratio > 300 mmHg under the condition in the hospital room (mild illness); 5. Willing to receive a random assignment to any designated treatment group and not participating in another study at the same time. The exclusion criteria: 1. Severe and critical illness patients or participating in the trial does not meet the patient’s maximum benefit or does not meet any criteria for safe follow-up in the protocol after a doctor’s evaluation; 2. Retinopathy and other retinal diseases; 3. Conduction block and other arrhythmias; 4. Severe liver disease (e.g., Child-Pugh score ≥ C or AST> twice the upper limit); 5. Pregnant or breastfeeding; 6. Severe renal failure [estimated glomerular filtration rate (eGFR) ≤ 30 mL/min/1.73m^2^] or receiving renal replacement therapy; 7. Possibility of being transferred to another hospital within 72 h; 8. Received any trial treatment for COVID-19 within 30 days before this research. 62 patients who met the trial criteria were randomly assigned in a to two groups, all received the standard treatment (oxygen therapy, antiviral agents, antibacterial agents, and immunoglobulin, with or without corticosteroids), patients in the HCQ treatment group received additional oral HCQ (hydroxychloroquine sulfate tablets, Shanghai Pharma) 400 mg/d (200 mg/bid) between days 1 and 5 (Figure 1), patients in the control group with the standard treatment only. Randomization was performed through a computer-generated list stratified by site. Treatments were assigned after confirming the correctness of the admission criteria. Neither the research performers nor the patients were aware of the treatment assignments.

**Figure 1:**
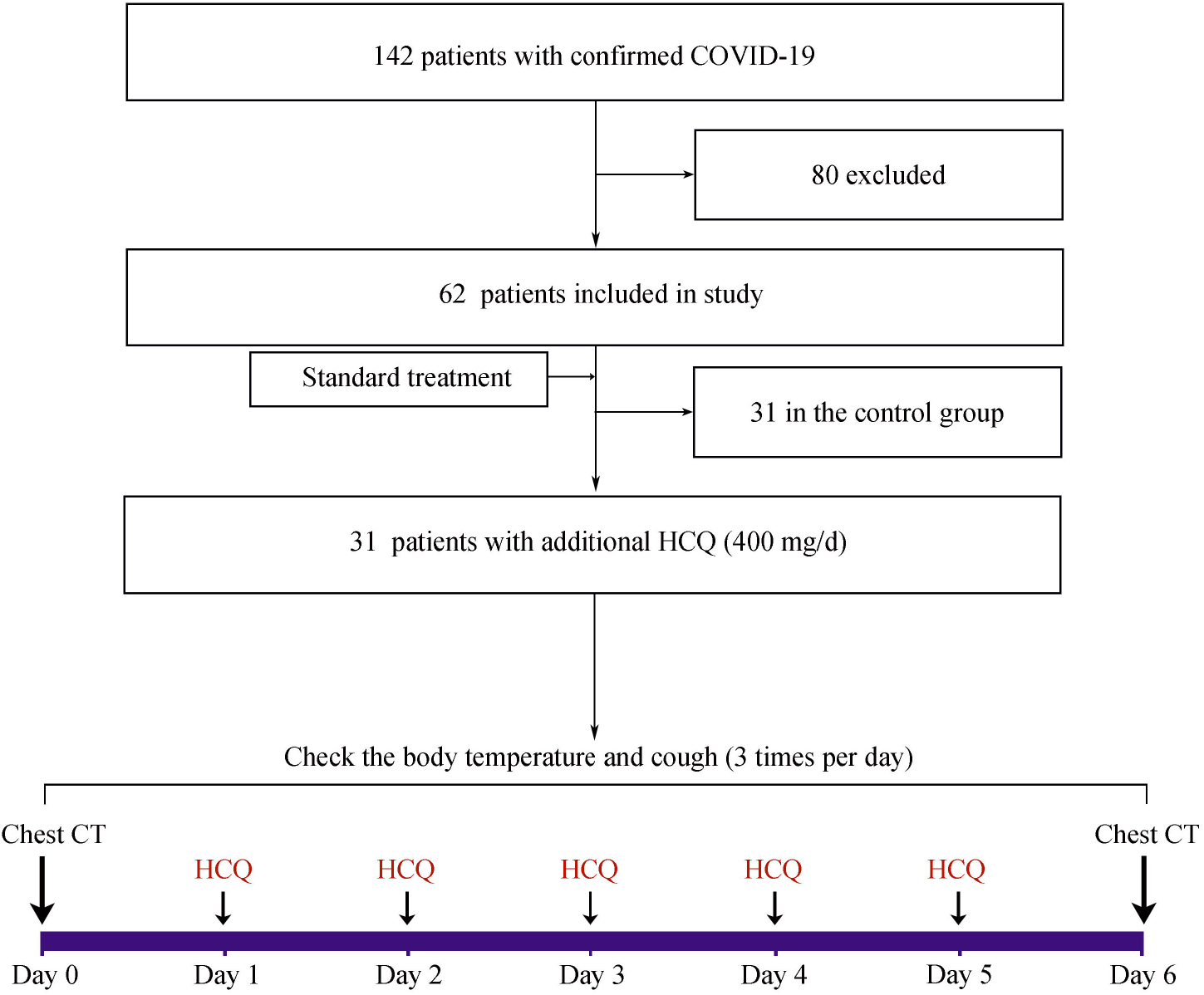
Study flow diagram. Abbreviations: COVID-19, severe acute respiratory syndrome coronavirus; HCQ, hydroxychloroquine; CT, computed tomography.

### Endpoints

5 days after enrollment or severe adverse reactions appeared was the observation endpoint. Changes in time to clinical recovery (TTCR) and clinical characteristics of patients were evaluated after administration. TTCR is defined as the return of body temperature and cough relief, maintained for more than 72 h. Normalization and mitigation criteria included the following: a. Body temperature ≤ 36.6 °C on the surface, ≤ 37.2 °C under the armpit and mouth or ≤ 37.8 °C in the rectum and tympanic membrane; b. Cough from patients’ reports, slight or no cough was in the asymptomatic range. Body temperature, cough check three times daily to calculate the average level. For radiological changes, the chest CT results in one day before (Day 0) and one day after (Day 6) the study for evaluation. Pulmonary recovery is defined as three levels: exacerbated, unchanged, and improved, moderately improved when less than 50 % of pneumonia were absorbed, and more than 50 % means significantly improved.

### Statistical Analysis

Data were described as the mean (standard deviation, SD), n (%), the t-test or χ^2^ test was used to compare the differences between the two groups. A two-sided p-value of less than 0.05 was considered statistically significant. Statistical analyses were performed using Graphpad Prism, version 6.0.

## Results

62 patients were identified as having COVID-19 and enrolled in this study, none quit (Figure 1). As shown in Table 1, For all patients, the age was 44.7 (15.3) years old, 46.8% (29 of 62) were male and 53.2% (33 of 62) were female. Patients were randomly assigned into two groups. There was no significant difference in the age and sex distribution between the two groups of patients, but there are significant differences in TTCR between the two groups. For fever, 17 patients in the control group and 22 patients in the HCQ treatment group had a fever in day 0. Compared with the control group [3.2 (1.3) days], the body temperature recovery time was significantly shortened in the HCQ treatment group [2.2 (0.4) days]. For cough, 15 patients in the control group and 22 patients in the HCQ treatment group had a cough in day 0, The cough remission time was significantly reduced in the HCQ treatment group. Notably, a total of 4 of the 62 patients progressed to severe illness, all of which occurred in the control group not receiving HCQ treatment. For adverse effects, it should be noted that there were two patients with mild adverse reactions in the HCQ treatment group, one patient developed a rash, and one patient experienced a headache, none severe side effects appeared among them.

**Table 1:** Characteristics of patients in this trial.

| Characteristics | All | Control | HCQ | P value |
| --- | --- | --- | --- | --- |
| Cases, n | 62 | 31 | 31 |  |
| Age, mean (SD) | 44.7 (15.3) | 45.2 (14.7) | 44.1 (16.1) | 0.8809 |
| Sex, n (%) |  |  |  | 0.7991 |
| Male | 29 (46.8%) | 15 (48.3%) | 14 (45.2%) |  |
| Female | 33 (53.2%) | 16 (51.7%) | 17 (54.9%) |  |
| Fever, day (SD)<sup>a</sup> | 2.6 (1.0) | 3.2 (1.3) | 2.2 (0.4) | 0.0008 |
| Cough, day (SD)<sup>b</sup> | 2.4 (1.1) | 3.1 (1.5) | 2.0 (0.2) | 0.0016 |
| Progressed to severe illness | 4 (6.5 %) | 4 (12.9 %) | 0 |  |
| Adverse effects | 2 (3.2 %) | 0 | 2 (6.4 %) |  |

<sup>a</sup>22 patients in the HCQ treatment group,17 patients in the control group with a fever one day before the intervention. <sup>b</sup>22 patients in the HCQ treatment group,15 patients in the control group with a cough one day before the intervention. Abbreviations: SD, standard deviation; HCQ, hydroxychloroquine; CT, computed tomography.

**Table 2:** Absorption of pneumonia on chest CT.

| Group | All | Exacerbated | Unchanged | Improved |  |  |
| --- | --- | --- | --- | --- | --- | --- |
|  |  |  |  | Moderate | Significant | Total |
| All | 62 | 11 (17.7 %) | 9 (14.5 %) | 18 (29.0 %) | 24 (38.7 %) | 42 (67.7 %) |
| Control, n (%) | 31 | 9 (29.0 %) | 5 (16.1%) | 12 (38.7 %) | 5 (16.1%) | 17 (54.8%) |
| HCQ, n (%) | 31 | 2 (6.5 %) | 4 (12.9 %) | 6 (19.4%) | 19 (61.3%) | 25 (80.6%) |
| P value | 0.0476 |  |  |  |  |  |

Abbreviations: HCQ, hydroxychloroquine.

To further explore the effect of HCQ on pneumonia, we compared and analyzed the chest CT of patients on day 0 and day 6. In our study, pneumonia was improved in 67.7% (42/62) of patients, with 29.0% moderately absorbed and 38.7% significantly improved. Surprisingly, a larger proportion of patients with improved pneumonia in the HCQ treatment group (80.6%, 25 of 31) compared with the control group (54.8%, 17 of 31). Besides, 61.3% of patients in the HCQ treatment group had a significant pneumonia absorption.

## Discussion

CQ and its derivatives have been broadly used as immunomodulators in the treatment of systemic lupus erythematosus (SLE) and other rheumatism [12]. As the pharmacological mechanism of CQ is further elucidated, its additional clinical applications, especially the antiviral activity, are also increasingly valued [13]. The efficiency of CQ has been proven in a variety of viruses, including human coronavirus [[14], [15], [16]]. Researchers have even reported both prophylactic and therapeutic advantages of CQ for SARS-CoV infection [17]. The severe acute respiratory syndrome caused by SARS-CoV-2 in several patients is quite similar to SARS-CoV in 2002 and is currently seriously threatening global health by triggering COVID-19. However, none specific drugs are available for the prevention or treatment of COVID-19.

Recently, Wang et al. identified that CQ could effectively inhibit the replication and spread of SARS-CoV-2 *in vitro* [7]. Experts and guides for COVID-19 in China have also recommended chloroquine phosphate superior to the treatment of SARS-CoV-2 infection [8,9]. To assess the safety and effects of CQ in patients with COVID-19, we registered this trial in ChiCTR and chose HCQ (the sulfate and phosphate salts of CQ) as the intervention agent. The data in this study revealed that after 5 days of HCQ treatment, the symptoms of patients with COVID-19 were significantly relieved, manifesting as shorten in the recovery time for cough and fever. At the same time, a larger proportion of patients with pulmonary inflammatory has been partially absorbed in the HCQ treatment group, indicating the immune modulation and anti-inflammatory properties of HCQ in non-malarial diseases [17]. At present, the multiple actions of HCQ such as regulation in pro-inflammatory cytokines [e.g. Tumor necrosis factor-α (TNF-α), interleukin-1 (IL-1), interleukin-1 (IL-6)], antioxidant activities, also promote it widely performed in rheumatic diseases such as SLE [11,17]. According to current research, a higher pro-inflammatory cytokine storm existed in COVID-19 patients with a severe or critical illness, eventually affected the prognosis [18]. For this, IL-6 antibody blocker, transfusion of convalescent plasma, and other therapies have been applied to counteract the cytokine storm [19,20]. Therefore, with antiviral and autoimmune regulation effects, HCQ should be a protector in SARS-CoV-2 infection. In the present study, the reduced risk of progression to severe illness in patients with HCQ treatment also explained the intervention effect of HCQ on the pathological process of the COVID-19.

Although HCQ has proven to be effective, with advantages of inexpensive and easily accessible, its potential detrimental effects in viral diseases must also be taken seriously. Retinopathy is one of the major adverse reactions of long-term therapy with HCQ [21]. Besides, patients with rheumatoid diseases treated with HCQ occasionally experience arrhythmias [22]. Other rare adverse reactions caused by HCQ include gastrointestinal reactions, cramps, liver dysfunction, itching, headache, dizziness, insomnia, peripheral neuropathy [13]. Fortunately, deciding on individual treatment plans scientifically, monitoring adverse reactions timely, to avoid overdose, short-term application of HCQ is relatively safe.

## Conclusion

Despite our small number of cases, the potential of HCQ in the treatment of COVID-19 has been partially confirmed. Considering that there is no better option at present, it is a promising practice to apply HCQ to COVID-19 under reasonable management. However, Large-scale clinical and basic research is still needed to clarify its specific mechanism and to continuously optimize the treatment plan.

## Data Availability

The dataset supporting the conclusions of this article is included within the article.

## Acknowledgments

Respectful thanks to all patients and their families enrolled in this study. Funding: This study was supported by the Epidemiological Study of COVID-19 Pneumonia to Science and Technology Department of Hubei Province (2020FCA005).

## Ethics approval and consent to participate

The clinical research protocol was reviewed and approved by the Ethics Committee in Renmin Hospital of Wuhan University (Wuhan, China). All research procedures adhered to the tenets of the Declaration of Helsinki. This trial has been registered in the Chinese Clinical Trial Registry, the unique identifier: ChiCTR2000029559.

## Author Contributions

ZZ designed this study and major in the clinical management of patients, data collection, data analysis, and writing of the first draft. ZC and JH improved the data analysis and finished the manuscript. SJ helped the clinical management of patients. SH had roles in data collection and interpretation. ZZ and DY had roles in data analysis and data interpretation, RZ and BH helped the data collection. All authors reviewed and approved the final version of the manuscript.

## Availability of data and materials

The dataset supporting the conclusions of this article is included within the article.

## References

1. Zhu N, Zhang D, Wang W, et al. A Novel Coronavirus from Patients with Pneumonia in China, 2019. New Engl J Med. 2020. DOI: 10.1056/NEJMoa2001017.

2. Gorbalenya AE, Baker SC, Baric RS, et al. Severe acute respiratory syndrome-related coronavirus: The species and its viruses – a statement of the Coronavirus Study Group. bioRxiv. 2020:2020-2022.

3. Li Q, Guan X, Wu P, et al. Early Transmission Dynamics in Wuhan, China, of Novel Coronavirus-Infected Pneumonia. The New England journal of medicine. 2020. DOI: https://doi.org/10.1101/2020.02.07.937862.

4. Chen N, Zhou M, Dong X, et al. Epidemiological and clinical characteristics of 99 cases of 2019 novel coronavirus pneumonia in Wuhan, China: a descriptive study. The Lancet. 2020;395(10223):507–513. DOI: 10.1016/S0140-6736(20)30211-7.

5. World Health Organization. COVID-2019 situation reports-59. https://www.who.int/docs/default-source/coronaviruse/situationreports/20200319-sitrep-59-covid-19.pdf?sfvrsn=c3dcdef9_2.

6. Wang C, Horby PW, Hayden FG, Gao GF. A novel coronavirus outbreak of global health concern. The Lancet. 2020;395(10223):470–473. DOI: 10.1016/S0140-6736(20)30185-9.

7. Wang M, Cao R, Zhang L, et al. Remdesivir and chloroquine effectively inhibit the recently emerged novel coronavirus (2019-nCoV) in vitro. Cell Res. 2020. DOI: 10.1038/s41422-020-0282-0.

8. J. Gao, Z. Tian, X. Yang. Breakthrough: chloroquine phosphate has shown apparent efficacy in treatment of COVID-19 associated pneumonia in clinical studies. Biosci Trends (2020 Feb 19), 10.5582/bst.2020.01047. DOI: 10.5582/bst.2020.01047.

9. Colson P, Rolain JM, Lagier JC, Brouqui P, Raoult D. Chloroquine and hydroxychloroquine as available weapons to fight COVID-19 [published online ahead of print, 2020 Mar 4]. Int J Antimicrob Agents. 2020;105932. DOI: 10.1016/j.ijantimicag.2020.105932.

10. The U.S. Food and Drug Administration (FDA). Coronavirus (COVID-19) Update: FDA Continues to Facilitate Development of Treatments. https://www.fda.gov/news-events/press-announcements/coronavirus-covid-19-update-fda-continues-facilitate-development-treatments.

11. Rainsford KD, Parke AL, Clifford-Rashotte M, Kean WF. Therapy and pharmacological properties of hydroxychloroquine and chloroquine in treatment of systemic lupus erythematosus, rheumatoid arthritis and related diseases. Inflammopharmacology. 2015;23(5):231–269. DOI: 10.1007/s10787-015-0239-y.

12. Gordon C, Amissah-Arthur MB, Gayed M, et al. The British Society for Rheumatology guideline for the management of systemic lupus erythematosus in adults. Rheumatology (Oxford). 2018;57(1): e1–e45. DOI: 10.1093/rheumatology/kex286.

13. Schrezenmeier E, Dorner T. Mechanisms of action of hydroxychloroquine and chloroquine:implications for rheumatology. Nat Rev Rheumatol. 2020. DOI: 10.1038/s41584-020-0372-x.

14. Akpovwa H. Chloroquine could be used for the treatment of filoviral infections and other viral infections that emerge or emerged from viruses requiring an acidic pH for infectivity. Cell Biochem Funct. 2016;34(4):191–196. DOI:10.1002/cbf.3182.

15. E. Keyaerts, L. Vijgen, P. Maes, J. Neyts, M.V. Ranst. In vitro inhibition of severe acute respiratory syndrome coronavirus by chloroquine. Biochem. Biophys. Res. Commun., 323 (2004), pp. 264–268. DOI: 10.1016/j.bbrc.2004.08.085.

16. Vincent, M.J., Bergeron, E., Benjannet, S. et al. Chloroquine is a potent inhibitor of SARS coronavirus infection and spread. Virol J 2, 69 (2005). DOI: 10.1186/1743-422X-2-69.

17. Al-Bari MA. Chloroquine analogues in drug discovery: new directions of uses, mechanisms of actions and toxic manifestations from malaria to multifarious diseases. J Antimicrob Chemother. 2015;70(6):1608–1621. DOI:10.1093/jac/dkv018.

18. J Liu, S Li, J Liu. et al. Longitudinal characteristics of lymphocyte responses and cytokine profiles in the peripheral blood of SARS-CoV-2 infected patients. medRxiv 2020.02.16.20023671; DOI: https://doi.org/10.1101/2020.02.16.20023671.

19. Conti P, Ronconi G, Caraffa A, et al. Induction of pro-inflammatory cytokines (IL-1 and IL-6) and lung inflammation by Coronavirus-19 (COVI-19 or SARS-CoV-2): anti-inflammatory strategies [published online ahead of print, 2020 Mar 14]. J Biol Regul Homeost Agents. 2020;34(2):1. DOI:10.23812/CONTI-E. DOI: 10.23812/CONTI-E.

20. Chen L, Xiong J, Bao L, Shi Y. Convalescent plasma as a potential therapy for COVID-19 [published online ahead of print, 2020 Feb 27]. Lancet Infect Dis. 2020; S1473-3099(20)30141-9. DOI:10.1016/S1473-3099(20)30141-9.

21. Proano Cinthia., Kimball Glenn P. Hydroxychloroquine Retinal Toxicity. N. Engl. J. Med., 380(17), e27. DOI:10.1056/NEJMicm1304542.

22. Radke JB, Kingery JM, Maakestad J, Krasowski MD. Diagnostic pitfalls and laboratory test interference after hydroxychloroquine intoxication: A case report. Toxicol Rep. 2019;6:1040–1046. Published 2019 Oct 7. DOI: 10.1016/j.toxrep.2019.10.006.

